# Cumulative survival profiling: a new PAP-based method for detecting heteroresistance in staphylococcal clinical isolates

**DOI:** 10.1101/2020.08.10.20148502

**Authors:** Ramzi A. Alsallaq, Tina H. Dao, Jason W. Rosch, Elisa Margolis

**Author notes:** Address correspondence to Ramzi Alsallaq.

## Abstract

The area under the population analysis profile (PAP) is used in the gold standard method for detecting heteroresistance in staphylococci. We tested the hypothesis that the initial inoculum strongly influences the area under the population analysis profile. We sought to interpret this dependence and develop a new metric that lacks this dependence to retrospectively detect heteroresistance to vancomycin in coagulase-negative staphylococcal (CoNS) isolates.

We tested our hypothesis on 20 PAPs from the heteroresistant positive control isolate (Mu3) and 7 PAPs from one CoNS isolate which is associated with poor clinical response. The area under the PAP depended linearly (p<0.001) on the initial inoculum. We interpreted the slope to be the cumulative survival under vancomycine concentration gradient. The statistical distribution of the cumulative survival for Mu3 and the CoNS isolate constituted the cumulative survival profiles for each. The profiles reflected spectrum of response under vancomycine gradient with the left-tail of CoNS isolate profile located near the median of Mu3 profile indicating the heteroresistance of the CoNS isolate and that the most resistant in the spectrum are likely to be associated with poor clinical response. We estimated that about two-third of the CoNS from unique participants are heteroresistant with 80% of heteroresistant isolates may be associated with a poor clinical response.

## Background

Vancomycin is the last-resort treatment for infections caused by multi-drug–resistant bacteria, such as methicillin-resistant *Staphylococcus aureus* (MRSA). However, therapeutic failures with vancomycin have been reported in cases where *in vitro* assays measuring the clinical minimum inhibitory concentration (MIC) indicated susceptibility to vancomycin. These infections are caused by heterogeneous vancomycin-intermediate staphylococci. In this context, a single isolate represents a majority of the susceptible population to vancomycin, whereas a tiny proportion of bacterial cells demonstrate resistance to vancomycin [1, 2]. There have been several cases of reduced susceptibility to vancomycin only a few decades after the first use of vancomycin. In 1996, a 64-year-old man, who acquired MRSA pneumonia after an operation for primary lung cancer, experienced failed treatment after an extensive course of vancomycin [3]. The causative MRSA strain, Mu3, was isolated from sputum and was categorized as heterogenous vancomycin-resistant *S. aureus* (hVISA). Since then, various methods of detection have shown vancomycin heteroresistance in other staphylococcal strains, including coagulase-negative staphylococci (CoNS) [4–10].

The presence of the resistant subpopulation in multi-resistant or opportunistic pathogens (e.g., *S. epidermidis* and *S. haemolyticus*) contributes to reduced efficacy of vancomycin, limiting treatment options, especially for patients undergoing chemotherapy for whom vancomycin is the last resort against bacterial infection. The resistant subpopulation is very difficult to detect and is often misclassified by the common clinical diagnostic tests. The Clinical and Laboratory Standards Institute methods, which use an inoculum of approximately 10^4^ CFU per assay, cannot detect the resistant subpopulation, which is present at a frequency of 10^−6^ or lower [11]. Therefore, detecting heteroresistance in clinical isolates with high accuracy is critical for clinical decision making.

The gold standard method for detecting heteroresistance to an antibiotic in staphylococcal clinical isolates [12–15] is the modified area under the curve of the population analysis profile (PAP-AUC) method described by Wootton et.al. [16], which uses the Mu3 strain as a positive-control comparator. In the PAP-AUC method, a prepared initial inoculum of the bacterial isolate is plated into different plates forming a gradient of antibiotic concentrations, then the bacterial density that survived the antibiotic at each concentration is recorded to construct the population analysis profile (PAP). A similar PAP is prepared for Mu3 in tandem of a batch of one [17] or several [16] isolates’ PAPs and used for comparison. Usually, the initial inoculum values varies widely, by as much as 1.5log10 [18–23], across the batch and differs from that of the Mu3. It has been reported that the area under the PAP curve is influenced by the initial inoculum [10]. Further, the use of a single Mu3 PAP to detect heteroresistance in a batch of isolates, hinders the necessary cross-validation which require replicas of Mu3 PAPs [16] and falsely presumes that a single Mu3 PAP is a representative of the broad spectrum that typically characterizes heteroresistance

Here, we used replica PAPs of positive controls to study the nature, the statistical significance and interpretation of the dependence of AUC on the initial inoculum. We employed a PAP-based method that lacks the influence of the initial inoculum, characterizes the broad spectrum of heteroresistance, and allows cross-validation to detect heteroresistance and heteroresistance that is associated with poor clinical response in retrospective cohort of CoNS isolates that was obtained from participants in whom CoNS central-line associated bloodstream infection (CLABSI) developed during treatment from leukemia.

## Methods

### Bacterial isolates

We retrospectively studied 74 isolates obtained with permission from all patients in whom coagulase-negative staphylococcal central-line bloodstream infection (CoNS-CLABSI) developed during treatment for leukemia at our institution between January 1, 2010, and March 28, 2016. The information regarding these isolates were published before [24].

### Population analysis profiles (PAPs)

Population analysis profiling was the basis for all heteroresistance-related detection in clinical isolates. PAP was performed according to standard preparation techniques for staphylococci [16, 25]. Each isolate was grown in 10 *mL* TSB overnight at 37°C. Overnight stationary-phase cultures were then serially diluted 1:100 in 1×PBS, ranging from 10^−2^ to 10^−8^, and 100 *μL* of each dilution was streaked on BHI agar plates containing 0, 1, 2, 3, 4, 6, or 8 *μg/mL* of vancomycin. Each isolate was inoculated onto duplicate plates at each dilution at each vancomycin concentration. After incubation for 48 hours at 37°C, colonies were counted to determine and plot the counts of bacteria in *CFU/mL* growing at each antibiotic concentration. The resulting PAPs were used in the analyses. The area under the actual colony-counts was used unless specified otherwise; for example the area under the log10 of the colony-counts was used for Wootton et al.’s method [16].

The isolates were analyzed in two batches, with 44 isolates (59%) that were analyzed at a later stage requiring addition of 0.2% yeast extract (catalog number 212750, BD Sciences) to the overnight culture as a growth supplement. Each batch run was accompanied by replicate runs of Mu3 strain as a positive control (3 and 7 replicas for the first and second patch, respectively, with growth supplement added to each of the 7 replicas’ overnight cultures). To determine the effect of the initial inoculum on the area under the PAP, we prospectively obtained 10 more PAPs for Mu3 and 7 PAPs for 1 CoNS-CLABSI isolate (CoNSB18) from 1:2 dilutions and 2x and 4x concentrations to create initial inoculum values spanning the range 10^7^ to 10^9^ *CFU/mL* for Mu3 and 10^8^ to 10^9^ *CFU/mL* for CoNSB18 for overnight cultures that otherwise were prepared and used to generate PAPs as described previously.

### Etest

Each clinical isolate and Mu3 were grown in 10*mL* of TSB overnight at 37°C and were then diluted 1:100 in fresh 10 *mL* TSB. The bacterial strains were incubated at 37 °C until OD at 600 nm 0.2 (McFarland unit 2). Then 100 *μL* of the culture were spread on BHI before placing the vancomycin Etest strips (bio-Mérieux, Marcy-l’Étoile, France) on the plate. The plates were incubated at 37 °C for 48 hours. Each isolate was classified as heteroresistant if isolated colonies were observed in the inhibition eclipse, as determined by two independent readings.

### Quantitative and statistical analysis

We created a new format for the analyses whereby each singlet PAP contributes a row, and the columns enlist the bacterial counts in *CFU/mL* for the PAP at 0,1,2,3,4,6, and 8 *μg/mL* of vancomycin. Each PAP from the isolates and Mu3 was first checked for missing initial inoculum or for having two data points or fewer and dropped if either was true. None of the 20 Mu3 PAPs were dropped, but 14 duplicated PAPs from 7 isolates and 3 singlets were dropped from the analyses for not passing either of the two filters. The remaining isolate PAPs were then divided into two unique isolate sets: training and verification sets of 66 and 65 singlet PAPs from 67 isolates, which included 56 and 54 first-episode isolates from 56 and 54 patients, respectively.

The area under the counts and the area under the log10 counts were calculated by using *auc* module from the Python package scikit-learn and were validated with corresponding calculations by using GraphPad Prism 7.

The linear relationship between the area under the bacterial counts and the counts at 0 *μg/mL* (the initial inoculum) was firstly explored using ordinary linear square regression fit; the goodness of which was judged by the p-values for the regression coefficients (slope and intercept) and the R-squared value. A better fit in terms of the p-values for the coefficients was obtained using mixed effect linear regression model with random intercept to adjust for PAP to PAP variation. For the cumulative survival profiling (CSP) method we obtained the distribution of the cumulative survival under vancomycin gradient of the reference strain using a mixed-effect linear regression analyses within Bayesian framework (further details are in the Supplementary Information Appendix). Because obtaining replica PAPs is not practical for the CoNS-CLABSI isolates, we estimated the cumulative survival using a single PAP by dividing the area under the counts by the initial inoculum. We call this estimate point-estimate-profiling (PEP).

We used the 2.5% quantile of the Bayesian credible interval of the distribution obtained by CSP for the reference as a threshold for detection. The detection accuracy of this threshold is determined through cross-validation over 50 iterations by dividing available PAPs of the reference into reference (80% of PAPs) and validation (20%) sets randomly selected in each iteration and then obtaining the mean and standard deviations of the percentage of validation PAPs detected by the method over the iterations. An isolate is heteroresistant if its estimated cumulative survival using PEP is ≥ the 2.5% quantile of the Bayesian credible interval of the cumulative survival estimated from the reference set.

The cross validation of heteroresistance detection over 20 Mu3 PAPs was compared for four methods: CSP, Wootton’s (AUC under the log10 counts) [16, 18, 21, 25, 26], semi-Wootton’s (AUC under the counts) [27–29] and altered semi-Wootton’s (counts are altered to match the initial inoculum of the reference before calculating AUC) [10]. The detection accuracy via cross-validation for each method was estimated like the CSP method except for the detection criterion. For all methods except the CSP, a given PAP from the validation set is heteroresistant if AUC is ≥ 0.9 the AUC of the averaged PAP of the reference.

Following successful cross-validation, credible detection of heteroresistance in the CoNS-CLABSI isolates by the CSP method is deemed possible. The total number of twenty Mu3 PAPs encompassed the reference set from which a distribution of the cumulative survival was obtained and the cumulative survival for each isolate was estimated by PEP and compared with the 2.5% quantile of the reference similar to what is described above for the Mu3 validation set.

To detect heteroresistance that may be associated with poor clinical response, we used CSP on 7 PAPs from 1 of the CoNS isolates (CoNSB18), which is found to be both heteroresistant and associated with poor clinical response under vancomycin treatment. As was done for Mu3, the detection accuracy was measured first by cross-validation. Association of detections with the measured poor clinical response was determined by using Cox proportional hazard regression.

We verified the accuracy and robustness of our CSP-based detections over replicated PAPs of isolates by comparing heteroresistance detection (with and without association with poor clinical response) for duplicates of 64 common isolates in the training and the verification sets.

## Results

### Initial inoculum strong influence on PAP-AUC

It has been reported by one study [10] that the initial inoculum has a large influence on PAP-AUC. Using 20 Mu3 PAPs, we confirmed the strong association between the area under the counts and the initial inoculum (*R*^*2*^ = 0.90 in the log space; Figure 1 A). A one log10 *μg/mL* change in the initial inoculum of Mu3 increases the area under the PAP counts by 1.0 log10 *CFU*/*mL*.*μg/mL* (p<0.001). This strong dependence holds true also for clinical isolates (*R*^*2*^ = 0.90; Figure 1 B) by plating one selected CoNS isolate (CoNSB18) using broad range of initial inoculum values. These results indicate that in PAP-based methods which detect heteroresistance by comparing PAP-AUCs, differences in the initial inoculums of Mu3, and an isolate such as CoNSB18 will propagate to influence their heteroresistance detection. An alternative metric to the area under the curve that is free from the influence of the initial inoculum is needed to adequately compare isolates to Mu3.

**Figure 1:**
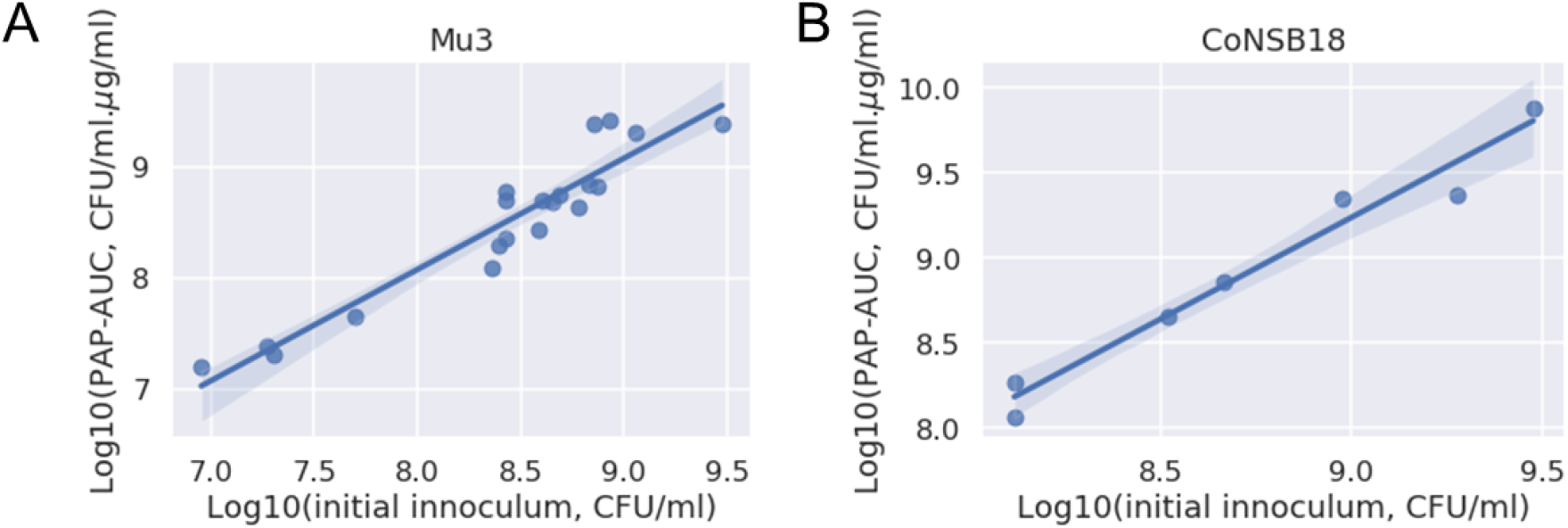
The strong dependence of the area under the counts on the initial inoculums for A) Mu3 using 20 PAPs and B) a selected CoNS isolate (CoNSB18) using 7 PAPs. In both cases, *R*^*2*^ = 0.90 and p<0.001.

### New metric: the cumulative survival under the antibiotic concentration gradient

The strong relationships in Figure 1 suggest the following model:

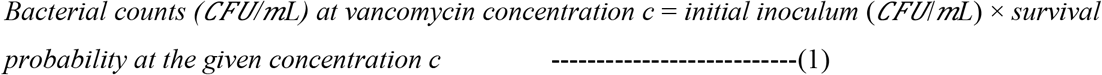

which leads to (see Supplementary Information Appendix for the derivation and explanation)

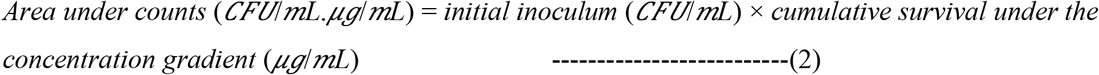

That is, for a single isolate (e.g. Mu3 or CoNSB18 in Figure 1), we interpret the y-intercept in Figure 1 as the log10 of the “cumulative survival under the antibiotic gradient”. In the above model, the information specifying the experience of the isolate under an antibiotic gradient is fully specified by the cumulative survival under that gradient, which is the needed metric free from the influence of the initial inoculum.

### Cumulative survival profiling (CSP) of Mu3

Here we used the CSP method to infer the full distribution of the cumulative survival of Mu3 under a vancomycin concentration gradient up to 8 *μg/mL* utilizing 20 Mu3 PAPs.

The Bayesian linear mixed-effects regression model fits data well, as shown in Figure 2 A, and the inferred cumulative survival for Mu3 (Figure 2 B) reveals not a single value but a wide distribution, with a median of 1.14 *μg/mL* and a 95% Bayesian credible interval (BCI) of 0.82-1.54 *μg/mL*. To utilize this distribution, we use the 2.5% quantile of the 95% BCI (q2.5%) to verify and detect heteroresistance of Mu3 profiles and of clinical isolates, respectively.

**Figure 2:**
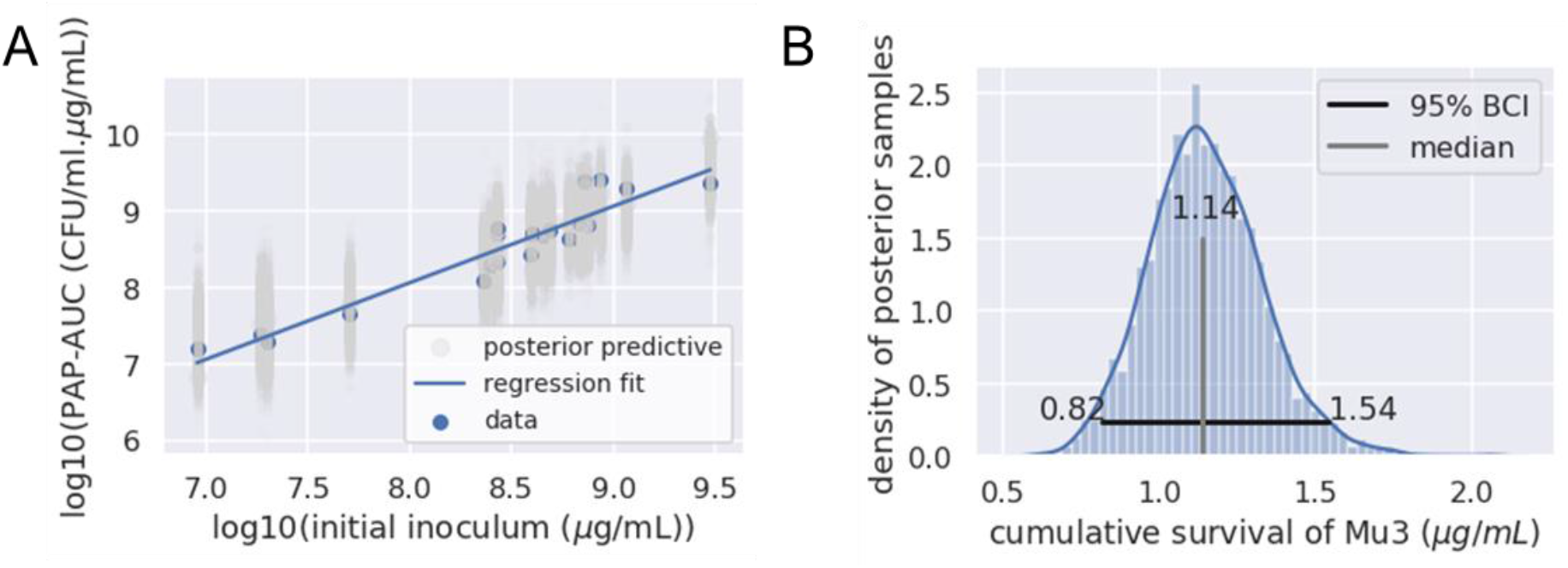
Cumulative survival profiling (CSP) of Mu3. A) Bayesian linear mixed-effects regression fit, B) the inferred distribution of the cumulative survival of Mu3 under a vancomycin gradient up to 8 *μg/mL*.

### Cross-validation of Mu3 heteroresistance: comparison with other methods

Compared to three other PAP-based methods used in the literature, the q2.5% of the inferred distribution of cumulative-survival of Mu3 (CSP method) was superior (Table 1), cross-validating the heteroresistance of Mu3 PAPs, on average, 82% of the time. This indicates that the CSP of Mu3 would detect heteroresistance in clinical isolates at a rate higher than that of the three other methods.

**Table 1:**
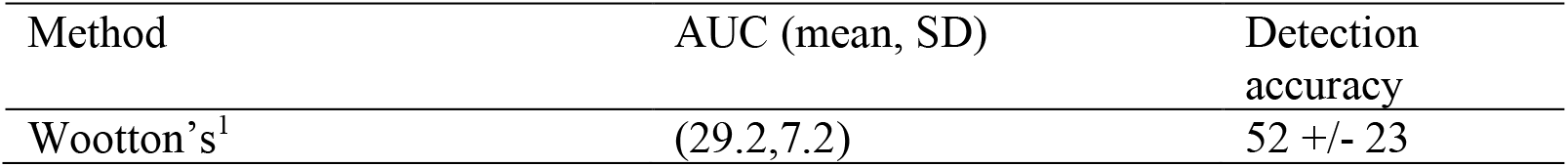
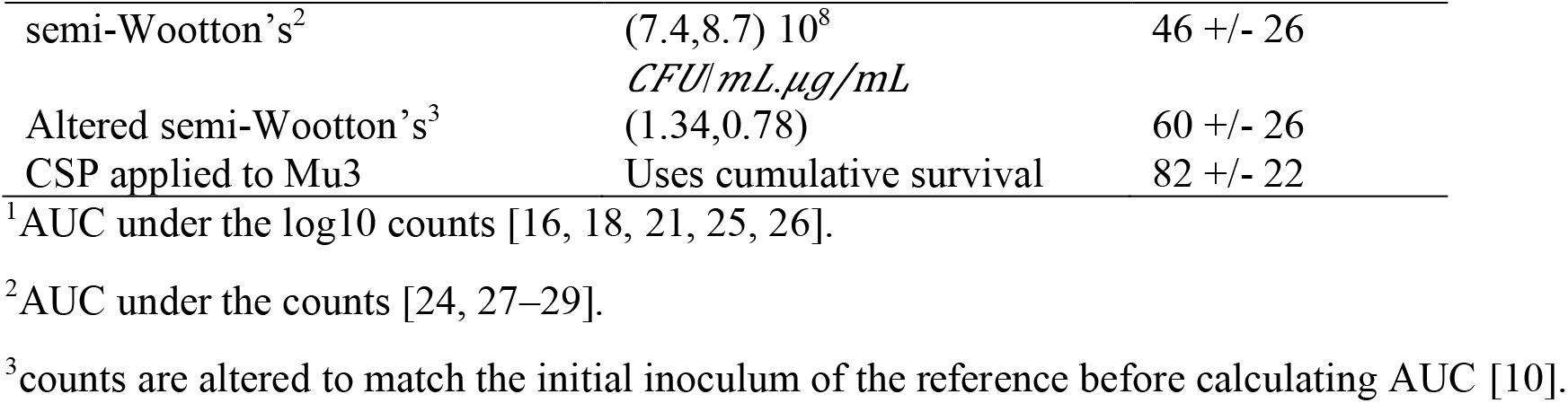
Cross-validation of 20 Mu3 PAPs using four methods. SD=standard deviation. Detection accuracy=% of Mu3 PAPs detected to be heteroresistant.

### Prevalence of heteroresistance in CoNS-CLABSI

Next, we used the cross-validated q2.5% threshold to detect heteroresistance in a set of 66 CoNS-CLABSI PAPs with 56 PAPs representing first-episode samples from 56 participants (training set). Heteroresistance was detected in 37, suggesting a 66% prevalence among participants from whom first-episode CoNS-CLABSI were sampled (Table 2).

**Table 2:** Prevalence of heteroresistance in training and verification sets of CoNS-CLABSI as revealed by q2.5% of the CSP of Mu3. The numbers and percentages of poor clinical response cases as measured by a clinical laboratory in our institution are also shown.

| CoNS-CLABSI set | Total |  |  | First episode |  |  |
| --- | --- | --- | --- | --- | --- | --- |
|  | N | Heteroresistant,<br>n (%) | Poor clinical response,<br>n (%) | N | Heteroresistant,<br>n (%) | Poor clinical response,<br>n (%) |
| Training | 66 | 46 (70) | 18 (27) | 56 | 37 (66) | 15 (27) |
| Verification | 65 | 48 (74) | 18 (28) | 54 | 39 (72) | 15 (28) |

We verified the accuracy of our detections over replicated PAPs of isolates by using another set of 65 CoNS-CLABSI PAPs (verification set) with 64 replicated PAPs of isolates in the training set and 54 PAPs representing the first episode from 54 participants. The estimated prevalence of heteroresistance in the first-episode CoNS-CLABSI PAPs in the verification set is 72% (Table 2). The CSP method accurately classified 58 of 64 replicas by heteroresistance status, giving an accuracy of 91% over replicated PAPs.

The association of detected heteroresistance with measured poor clinical response for CoNS-CLABSI is not statistically significant (p>0.05) regardless of the set of isolates. This is likely because, within the spectrum of the hVISA heteroresistance revealed by the distribution for Mu3 in Figure 2 B, only those with the highest cumulative survival might be intuitively associated with poor clinical response.

### Detection of poor clinical response attributable to heteroresistant CoNS-CLABSI

To establish whether poor clinical response might be attributable to detected heteroresistance, we needed to estimate the proportion of CoNS-CLABSI that are *potentially* associated with poor clinical response by profiling a CoNS-CLABSI isolate that is both heteroresistant and associated with poor clinical response. The isolate CoNSB18, with the PAP replicas in Figure 1B, fitted these characteristics. The CSP for this isolate revealed a cumulative survival of 1.76 (95% BCI:1.15-2.81) *μg/mL*, suggesting a threshold of 1.15 *μg/mL* to detect heteroresistant CoNS-CLABSI that may be associated with poor clinical response. Cross-validating the 7 PAPs for this isolate by using this q2.5% threshold gave an average detection accuracy of 80% (SD=33%) for the heteroresistance that may be associated with poor clinical response.

We then detected similar isolates among our CoNS-CLABSI samples using this cross-validated threshold (Table 3). Cox proportional hazard regression analyses reveal statistically significant association between our detections and time to measured poor clinical response or censorship (p<0.05), regardless of the set of isolates (Table 3). We estimated that more than 80% (31/37 and 32/39 from the training and verification sets, respectively) of heteroresistant CoNS-CLABSI may be associated with poor clinical response. The threshold that successfully detected heteroresistance with potential association with poor clinical response is about the median for Mu3 CSP (Figure 3).

**Table 3:**
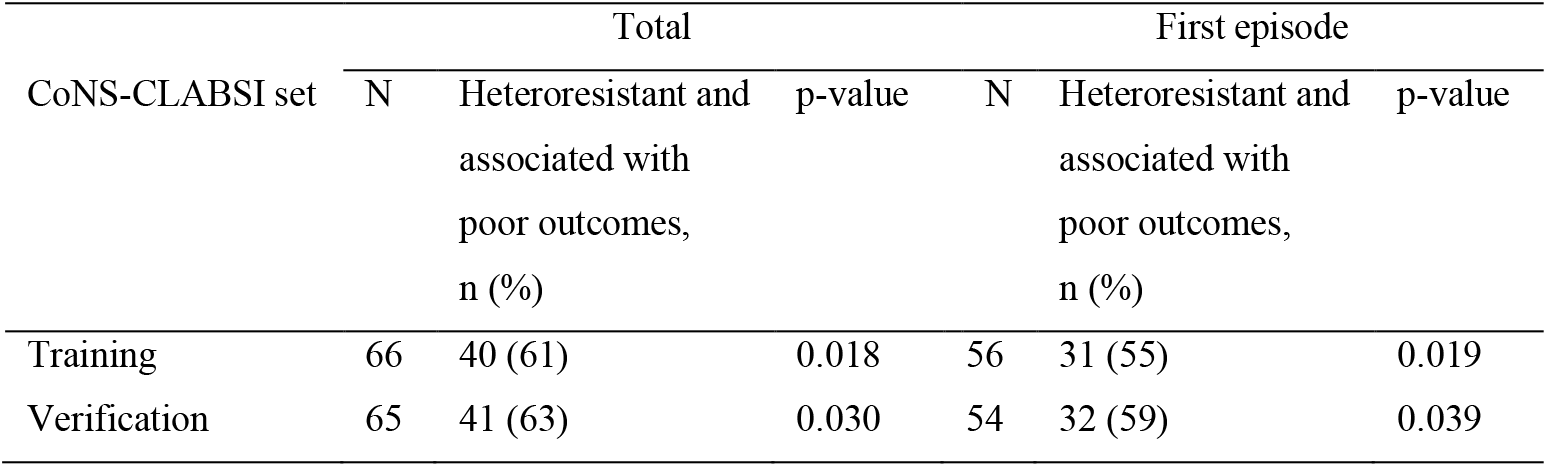
Prevalence of heteroresistance that is potentially associated with poor clinical response in training and verification sets of CoNS-CLABSI as revealed by q2.5% of the CSP of CoNSB18.

| CoNS-CLABSI set | Total |  |  | First episode |  |  |
| --- | --- | --- | --- | --- | --- | --- |
|  | N | Heteroresistant and associated with poor outcomes, n (%) | p-value | N | Heteroresistant and associated with poor outcomes, n (%) | p-value |
| Training | 66 | 40 (61) | 0.018 | 56 | 31 (55) | 0.019 |
| Verification | 65 | 41 (63) | 0.030 | 54 | 32 (59) | 0.039 |

**Figure 3:**
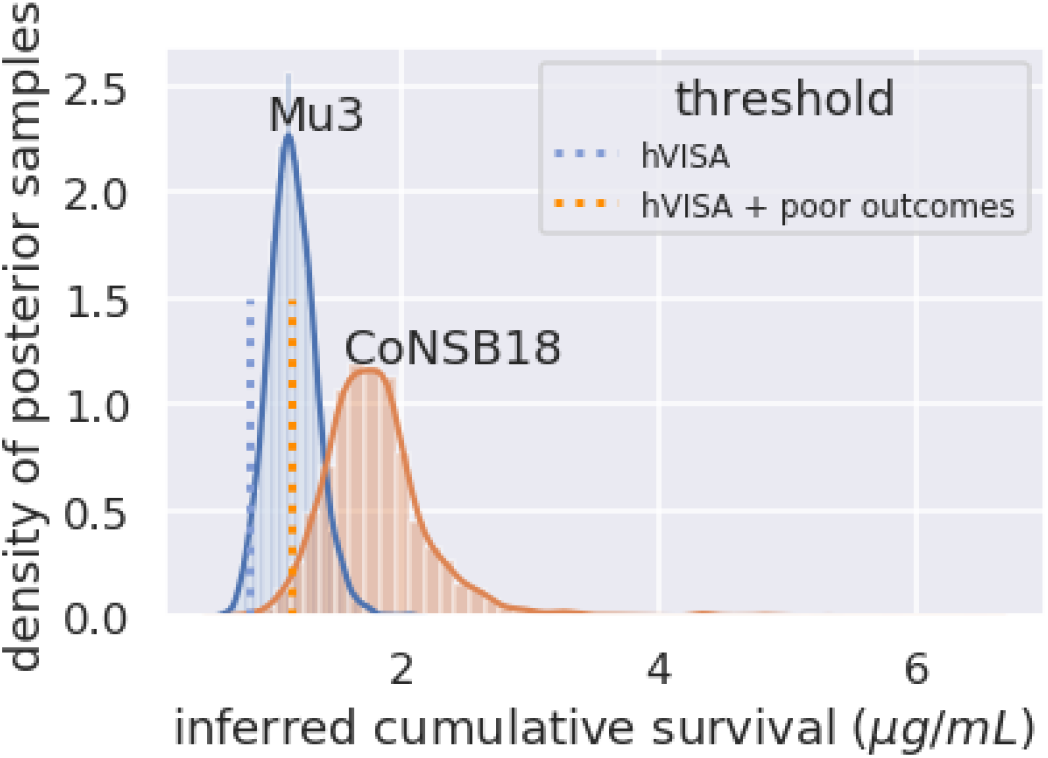
Inferred cumulative survival profiles for Mu3 and CoNSB18. The cumulative survival threshold that successfully detected heteroresistant isolates with potential association with poor clinical response (hVISA + poor outcomes) at 1.15 *μg/mL* is higher than the threshold that successfully detected all heteroresistant isolates regardless of association with clinical outcomes (hVISA) at 0.82 *μg/mL*.

### Comparison with Etest

Next, we compared Etest classification results to the two heteroresistance classifications: Mu3-like and CoNSB18-like detection, taking each separately as the gold standard. Etest accurately predicted the heteroresistance status of 60% and 62% of the isolates in the training set as respectively classified by Mu3-like and CoNSB18-like detection. The corresponding sensitivities of Etest were 75% and 79% and the specificities were 26 and 33%, respectively. The results were robust across PAP replicas (Table 4).

**Table 4:**
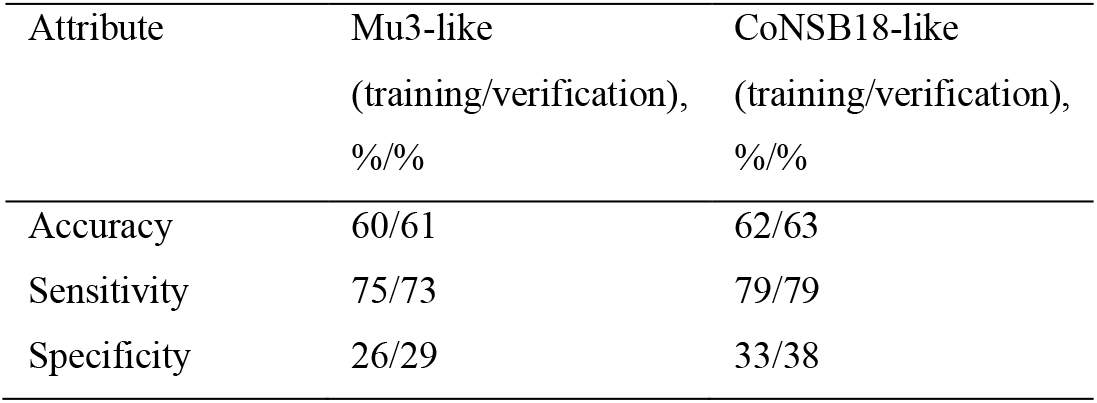
Comparing Etest classification to the classifications by CSP of Mu3 and of CoNSB18, taking each separately as the gold standard.

| Attribute | Mu3-like<br>(training/verification),<br>%/% | CoNSB18-like<br>(training/verification),<br>%/% |
| --- | --- | --- |
| Accuracy | 60/61 | 62/63 |
| Sensitivity | 75/73 | 79/79 |
| Specificity | 26/29 | 33/38 |

## Discussion

In this study of heteroresistance detection among hospital 74 CoNS isolates we have shown that the initial inoculum used in PAP-based heteroresistance detections in staphylococci statistically significantly influences (p<0.001) the area under the PAP curve. This finding implies that the area under the PAP curve is not adequate for evaluating the experience under the antibiotic gradient. We introduced the cumulative survival under the antibiotic gradient as an alternative metric and a distribution of which must be estimated for reference strains. To infer the distribution of the cumulative survival for reference strains, we introduced the cumulative survival profiling (CSP) method which employs replicated PAPs of the reference with a wide range of initial inoculum values. For clinical isolates, that one aims to compare with reference strains, the cumulative survival can be estimated, simply, by dividing the area under the PAP curve by the initial inoculum. We applied the CSP method on two reference isolates Mu3 (heteroresistant) and CoNSB18 which is detected to be heteroresistant in comparison with Mu3 and is associated with poor clinical response. Using these two reference strains, we estimated that about two-thirds of our first-episode CoNS-CLABSI isolates are heteroresistant to vancomycin and that 80% of the heteroresistant isolates may be associated with a poor clinical response. These estimates raise the profile of heteroresistance to vancomycin as a risk factor for poor clinical response in CoNS-CLABSI isolated from the pediatric leukemia population.

### Comparison with other studies

Our estimate of the heteroresistance prevalence in CoNS under vancomycine is consistent with the 68% prevalence of heteroresistance among MRSA CoNS [5] and with the 62.5% prevalence estimate among MRSA *S. captis* from blood samples in neonatal intensive care units (NICUs) [8] and within the range reported among CoNS in NICUs [7] but it is higher than what we estimated for the same data using the semi-Wotton’s method [24] because the latter revealed heteroresistance 46% of the time compared to 82% of the time using the CSP method.

### Strength and limitations of this study

The inferred cumulative survival distributions of Mu3 and of CoNSB18 under vancomycin reveal the wide spectrum of heteroresistance and that a single PAP would not be sufficient to capture. Further, the finding that a larger cumulative survival threshold (median compared to 2.5% quantile of the CSP of Mu3) was needed to successfully detect heteroresistance associated with a poor clinical response than that needed to detect heteroresistance regardless to association with clinical outcomes shows that only the most-resistant CoNS isolates (those with relatively higher cumulative survival above the median CSP distribution of Mu3) may be associated with poor clinical response.

The CSP method offers asynchronous profiling of Mu3 and prospective isolates, whereby an experimental setup is used to first conduct PAPs for Mu3 and build its cumulative survival profile. The detection accuracy of heteroresistance by the Mu3 profile is then examined via cross-validation over the Mu3 PAPs. More Mu3 PAPs can be added overtime to this profiling provided following the same experimental setup in preparation and the re-examination of the detection accuracy. The exact same experimental setup is then used to conduct PAPs for isolates as they come forward and the most up-to-date Mu3 profile is used to detect heteroresistance in each. Thus, this method alleviates the need for profiling clinical isolates simultaneously with the reference strains and could pave the way to standardization of the detection of heteroresistance.

Along these lines, the asynchronous profiling must not mean different experimental procedures are used for the reference and isolates’ PAPs. Any differences should be investigated and reported. In this regard, the CSP method explicitly controls for the variation in the initial inoculum and only the random experimental errors occurring within the same experimental procedure.

Also, the CSP method makes it feasible to detect strains that are not only heteroresistant but also associated with poor clinical response by profiling an isolate that is known to be associated with poor clinical response provided it is found to be heteroresistant in comparison with Mu3 profile. This isolate then can be used as a reference to detect similar isolates.

By including cross-validation, the CSP method ensures the reliability of reference-like detection in new clinical isolates. The cross-validation of the CSP method reveals a false-negative rate of <20%. Although this false-negative rate might never by zero for Mu3 given that some PAPs of Mu3 inherently reflect susceptibility to vancomycin (which again argues against using single Mu3 PAPs for detecting heteroresistance), using more PAPs for the reference would decrease this rate.

Our CSP profiles shown in this study are specific to the vancomycin gradient and might be specific to the experimental preparation that was followed for PAPs and should not be directly applicable to other antibiotics, different vancomycin gradients, or different experimental preparations. Apart from Mu3, we have not tested our method on *S. aureus* isolates, and the samples of CoNS used might not represent other populations. Using different methods, it has been observed that prevalence of hVISA could vary among populations [27]. This variability suggests that an adequate reference isolate is always needed to predict/confirm association with clinical outcomes of prospective isolates.

Therefore, we are making the method publicly available at (https://github.com/stjude/CSP) so that other investigators may test it on their isolates prospectively or retrospectively.

Although the most robust estimate for the cumulative survival would require profiling each clinical isolate by applying CSP using multiple PAPs of the isolate, in case of multiple clinical isolates, a single PAP for each might be enough. However, this supposition warrants further testing. We showed robustness of classification results to replicated PAPs of isolates.

The comparison of detection results of Etest to CSP-detection of CoNSB18 indicate a relatively low false-negative rate of 20%, suggesting that Etest might be used first on CoNS to detect hVISA-like heteroresistance associated with poor clinical response, with an understanding of the limitation of this false-negative rate.

### Conclusions and clinical implications

In summary, the initial inoculum used in PAP-based heteroresistance detections in staphylococci strongly influences the area under the PAP curve and should not be part of evaluating the experience of isolates under the antibiotic gradient. We suggest using the cumulative survival instead. The cumulative survival profiling of Mu3 using multiple PAPs reveals a spectrum for the experience of Mu3 under vancomycin gradient indicating that robust determination of heteroresistance in clinical isolates using Mu3 cannot be adequately captured by a single Mu3 PAP.

## Data Availability

data is available on the link below

https://github.com/stjude/CSP

## Acknowledgment

We thank Elaine Tuomanen for discussion and encouragement. We thank Cherise Guess, PhD ELS for scientifically editing the manuscript.

## Funding

EM receives funding from ALSAC. JWR is supported by 1U01AI124302 and 1RO1AI110618.

## Notes

Competing interest: All authors declare no competing or conflict of interest

### Competing Interest Statement

The authors have declared no competing interest.

### Author Declarations

The study was approved by our institution review board (XPD16-036)

