## Supplementary Information Appendix for "Cumulative survival profiling: a new PAP-based method for detecting heteroresistance in staphylococcal clinical isolates"

### Data Availability

The data and the codes are available at https://github.com/stjude/CSP

### Deriving the cumulative survival under vancomycin from PAP profile

Recasting Eq.1 in the main text using mathematical symbols for convenience:

$$B(c) = CFU_0 \times S(c) \quad \text{-------------------------------------} \ \text{(S1)}$$

Where ($CFU_0$) is the initial inoculum and does not depend on the concentration of antibiotic ($c$) and $S(c)$ is the concentration-dependent survival fraction.

The cumulative survival can be derived from the PAP by mathematically integrating Eq. S1 over vancomycin gradient:

$$\int_0^{c_{max}} B(c)dc = \int_0^{c_{max}} CFU_0 \times S(c)dc = CFU_0 \times CumSurvival \quad \text{-------------------------}\text{(S2)}$$

The left-hand side of Eq.S2 is the area under the counts. Thus, the cumulative survival can be obtained from a single PAP by dividing the area under the counts by the initial inoculum and for multiple PAPs by using regression analysis.

### Bayesian mixed-effects regression analysis

For inferring the cumulative survival distribution for reference strains in the CSP method, we used Bayesian linear mixed-effects model. To do that we have $1,2,3,\ldots,N$ PAPs that span a wide range of initial inoculum. Let the areas under the counts for these PAPs in $CFU/mL.\mu g/mL$ be $y_i$ and the initial inoculums be $x_i$. We wish to infer the cumulative survival $\xi$. From Eq.S2 we know that:

$$log10(y_i) = log10(x_i) + log10(\xi) \quad \text{-------------------------}\text{(S3)}$$

To incorporate the random effect representing PAP to PAP variation, we introduce zero-centered random deviations from the mean initial inoculum of each PAP. Let this deviation be $\epsilon_i \sim N(0,10^8)$, then the mixed-effects regression model becomes:

$$log10(y_i) = log10(x_i + \epsilon_i) + log10(\xi) \quad \text{-------------------------}\text{(S4)}$$

Equation S4 defines the mean of our model and can be fitted with zero-centered residuals:

$$log10(y_i) \sim N(log10(x_i + \epsilon_i) + log10(\xi), \sigma^2) \quad \text{-------------------------}\text{(S5)}$$

Posterior samples were obtained by using MCMC with NUTS sampler by utilizing the python package PYMC3 [1]
